# PATTERNS OF ANAESTHESIA(AS) AND ANALGESIA(AG) PRACTICE IN GYNECOLOGICAL BRACHYTHERAPY: THE INDIAN SCENARIO

**DOI:** 10.1101/2025.09.10.25335515

**Authors:** Pratiksha Tyagi, Amanda Rivera, Santam Chankraborty, Tapesh Bhattacharya

## Abstract

**INTRODUCTION:** Fractionated HDR brachytherapy, following external beam radiotherapy, is essential for treating locally advanced cervical cancer. Modern techniques enable delivering multiple fractions with a single application. However, applicator insertion, duration of it in situ and removal causes significant pain and distress. With no standardized guidelines for anesthesia in gynecological brachytherapy, this study surveyed current practices of anesthesia used across India to better understand and improve these issues.

**MATERIALS AND METHODS:** A survey was conducted with an online questionnaire that was shared with radiation oncologists through social media platforms (survey instrument: Rivera et al 2024). Analysis was done using descriptive statistics.

**RESULTS:** 93 responses were received, excluding 14 repeats 79 responses were analysed. Most of the respondents were male (52%). The most common place for intracavitary application is a dedicated brachytherapy suite. Spinal anesthesia, alone or in combination, was the most frequently used technique for intracavitary (39%), hybrid (30%), and interstitial (36%) brachytherapy. Most institutes are doing >20 applications per month. During waiting periods and treatments, oral analgesia is most commonly used. Nearly half (49%) of respondents perceived the procedure as distressing, yet fewer than half (44%) reported implementing measures to reduce psychological distress. Compared with Western data, Indian practice is characterized by higher procedure volumes, greater reliance on regional anesthesia, and limited use of conscious sedation.

**CONCLUSION:** We observed clinician preference for using the type of AG which highlights the Clinician bias in pain control in gynaecological brachytherapy. AG/AS during waiting times and applicator removal are areas of interest and shall be explored more. There is a need for good evidence for better management of pain and distress associated with gynaecological brachytherapy.

## Introduction

Cervical cancer is the leading cancer among women globally with the age standardised incidence rate (ASR) being 14.1/100,000 and ASR in India being 17.7/100,000 as per data in 2022. Notably, India was one of the top 3 countries with highest cervical cancer deaths in 2022.^1^ HDR brachytherapy is the standard of care for curative treatment of locoregionally advanced cervical cancer (FIGO stages IB2 to IVA) following concurrent chemoradiation. It can be intracavitary or interstitial with CT or MRI guidance.^2 3^ The pain that patients experience starts with applicator insertion, movement while shifting the patient and applicator in situ for 24-48 hours while the patient remains motionless in bed. The applicator in the uterus stimulates sympathetic autonomic nerve roots which enter the cord at T10-L1 level, which leads to cramping lower abdominal pain with nausea and vomiting. Distention and dilatation of the cervix and vagina and vaginal packing stimulates parasympathetic autonomic nerve roots and somatic nerve roots at S2-S4 level which leads to backache.^4^With pain, patients also experience emotional distress and anxiety.^6^Although several standardized guidelines exist for the management of the disease, there are no specific recommendations outlined for anaesthesia and analgesia for intracavitary or interstitial brachytherapy in cervical cancer. Limited data is available on the current patterns of practice in India. There have been surveys in the United States and the UK wherein the physicians, physicists or the specialist nurses have answered questionnaires regarding the type of pain reducing/analgesic/anaesthetic technique used during the application, treatment and applicator removal. ^7 13^Multiple studies confirm the psychological burden associated with brachytherapy. It has been observed that during brachytherapy treatment, patients experience elevated anxiety levels, exacerbated by inadequate communication and body exposure.^6 12 13^ Women describe the experience as distressing, with long-term emotional impact.^5 15 17^ Systematic reviews and qualitative studies consistently highlight the need for pre-treatment counselling, privacy measures, and continuity of care to mitigate psychological trauma.^15 16 17^

To understand the current scenario of patterns of practice of brachytherapy procedures and the type of analgesia used in India better, we conducted this survey wherein we studied the variable practices in anaesthesia used during the procedure across the country by radiation oncologists working at different centres. And also the physician’s perceived rates of patients’ emotional distress were evaluated. The aim of the study was to recognise the variability, knowledge gap, resource constraints and to be able to raise the concern for the need of a standard protocol for analgesia(AG) and anesthesia(AS) in gynaecological brachytherapy.

## Methods

The survey instrument used in this study has been previously used in the study by Rivera et al.^7^ Prior permission was obtained from the author before using it in our study. The questionnaire consisted of 31 questions that included basic demographic details of the institute, the age/gender of the respondent, and the gynaecological brachytherapy burden, practices, and AG/AS used. Specific questions about the race and ethnicity of the respondents were removed as they did not hold relevance to our setting.

An electronic version of the survey was created on Google Survey. The link was shared with radiation oncologists through social media platforms, and emails were sent to the NCG-registered institutes. QR codes for the survey were shared in various National conferences. A convenience sampling strategy was used. Additionally, email invitations were sent to the contacts of 260 National Cancer Grid-affiliated centres. These contact details were manually obtained from the website of the respective centres where available. The survey was conducted over a period of three months (September 2024 - November 2024). Only a single survey response per institute was accepted. Incomplete responses were discarded; however, if a single response was obtained for an institute, any incomplete response was retained for analysis. In cases where more than one response was provided for the type of anaesthesia, the highest level was used. (Considering the order of rank from highest to lowest as General anaesthesia, Conscious sedation, Spinal/Epidural anaesthesia, Oral analgesics). Descriptive statistics were used to analyse the data. Univariate descriptive statistics were used to determine the frequencies and percentages of various variables. Bivariate analysis was used to assess the association of anaesthesia type with other variables. Chi-square test or Fisher’s exact test was used to evaluate the statistical significance of the association between each categorical variable at a 0.05 significance level.

## Results

We received 93 responses from institutes across the country. A total of 49 responses were received from NCG-affiliated centers (Map 1). As the survey sampling strategy was not based on a defined sampling frame, calculation of the actual response rate is difficult. However, at the time of the survey, about 372 centres were affiliated with the National Cancer Grid. Of 93 responses, 14 were excluded as they were multiple responses from the same institute. A total of 79 responses were evaluated.

**Map 1.**
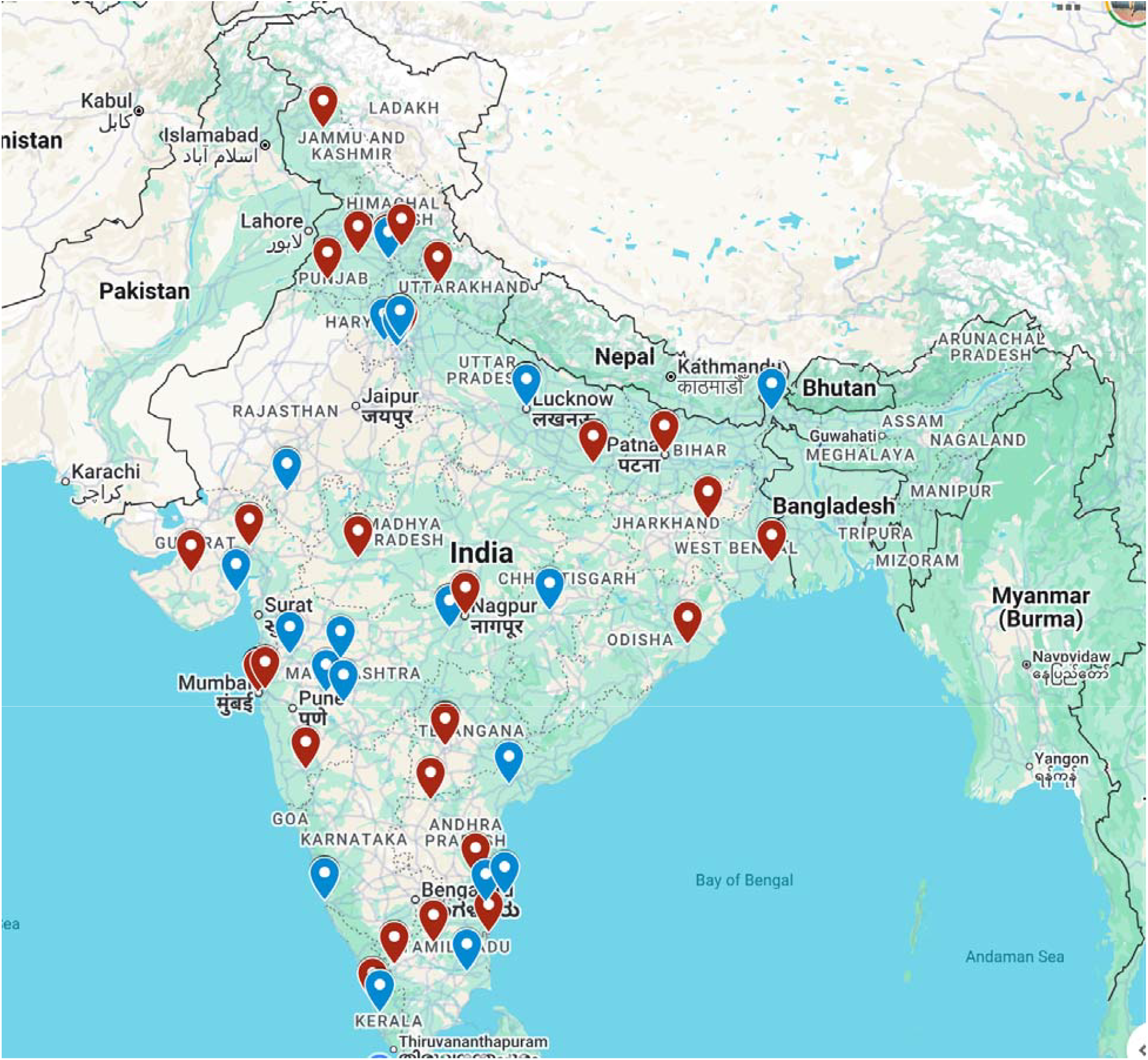
Showing all respondents (Blue) and NCG registered centers (Maroon)

### Demographics

Of 79 respondents, 52% were males. 58% were aged between 31-40 years. Most of the respondents were working in the centres located in the north and the south of India, followed by those in the western part of India. All respondents were radiation oncologists. (Table 1)

**Table 1.** Demographic characteristics of participants (N = 79)

| Characteristic | n (%) |
| --- | --- |
| Gender |  |
| Male | 41 (52%) |
| Female | 38 (48%) |
| Location |  |
| North | 23 (31%) |
| East | 11 (15%) |
| West | 15 (20%) |
| South | 24 (32%) |
| Northeast | 1 (1.4%) |
| Age group (years) |  |
| 25–30 | 16 (20%) |
| 31–40 | 46 (58%) |
| 41–50 | 11 (14%) |
| 51–60 | 6 (7.6%) |

### Gynaecological brachytherapy practices (Table 2)

Among the respondents, 37% (35/95) carry out the procedure in a dedicated brachytherapy suite. Note that some participants 5/79, performed brachytherapy in different areas in the same institute. 51% (48/95) had access to a dedicated OT where the procedure is carried out and 7.4% (7/95) performed the procedure in the CT simulator. Only 5/95 (5.3%) of the respondents performed insertions in the clinic examination room. Of the centres that do it in the clinical examination room, the majority used conscious sedation for application. During the waiting hours intravenous analgesics are used occasionally.

**Table 2.** Procedural and Technical Characteristics of Brachytherapy Practice (N = 79)

| **Applications per month** |  |
| --- | --- |
| 0–5 | 10 (13%) |
| 5–10 | 12 (15%) |
| 10–15 | 16 (20%) |
| 15–20 | 10 (13%) |
| >20 | 29 (37%) |
| N/A | 2 (2.5%) |
| **Admission for Brachytherapy** |  |
| No | 21 (27%) |
| Yes, routinely for intracavitary brachytherapy | 12 (15%) |
| Yes, routinely for intracavitary/interstitial brachytherapy | 17 (22%) |
| Yes, routinely for both | 29 (37%) |
| **Brachytherapy Location** |  |
| Brachytherapy Suite | 26 (33%) |
| Clinic exam room | 3 (3.8%) |
| CT sim | 2 (2.5%) |
| CT sim, Operating Room | 1 (1.3%) |
| CT sim, Operating Room, Brachytherapy Suite | 2 (2.5%) |
| CT sim, Operating Room, Clinic exam room, Brachytherapy Suite | 2 (2.5%) |
| Operating Room | 38 (48%) |
| Operating Room, Brachytherapy Suite | 5 (6.3%) |
| **USG guidance during application** |  |
| >80% | 33 (42%) |
| 30–70% | 11 (14%) |
| 10–30% | 19 (24%) |
| Never | 16 (20%) |
| **Type of USG guidance** |  |
| Transabdominal | 45 (57%) |
| Transrectal | 4 (5.1%) |
| Both | 11 (14%) |
| N/A | 19 (24%) |
| **CT guidance during application** |  |
| >80% | 5 (6.3%) |
| 30–70% | 3 (3.8%) |
| 10–30% | 11 (14%) |
| Never | 60 (76%) |
| **MRI guidance during application** |  |
| 30–70% | 2 (2.5%) |
| 10–30% | 10 (13%) |
| Never | 67 (85%) |
| **CT to confirm application placement** |  |
| >80% | 71 (91%) |
| 30–70% | 4 (5.1%) |
| 10–30% | 1 (1.3%) |
| Never | 2 (2.6%) |
| Unknown | 1 (1.3%) |
| **MRI to confirm application placement** |  |
| >80% | 1 (1.3%) |
| 30–70% | 8 (10%) |
| 10–30% | 15 (19%) |
| **Dose prescription in ICBT** |  |
| ICRU points prescription (point A, B etc) | 24 (30%) |
| HR-CTV prescription per GEC ESTRO guidelines | 34 (43%) |
| Both | 19 (24%) |
| N/A | 1 (1.3%) |
| **Hybrid application** |  |
| Yes | 44 (56%) |
| No | 35 (44%) |
| **Hybrid applicator used** |  |
| a. Vendor made (ie Elekta, Varian) | 28 (36%) |
| a. Vendor made (ie Elekta, Varian), b. Free hand | 6 (7.7%) |
| a. Vendor made (ie Elekta, Varian), b. Free hand, c. Institution made | 2 (2.6%) |
| Unknown | 2 (2.6%) |
| a. Vendor made (ie Elekta, Varian), c. Institution made | 4 (5.1%) |
| b. Free hand | 2 (2.5%) |
| c. Institution made | 1 (1.3%) |
| d. N/A | 36 (45.5%) |

67% (53/79) of the institutes performed 3 applications for a standard intracavitary brachytherapy procedure while 14% (11/79) used 4 applications; the remaining centres did 1-2 applications per patient. When a separate application was done for every fraction, there is a chance of an increase in pain distress experienced. 1 center that does 4 applications/patients reported using oral analgesia also.

Of the centers that did 4 applications for a patient, the majority did it in a brachytherapy suite or operating room and used spinal anaesthesia 46% (6/13)for intracavitary application.

Most of the respondents 37% (29/79) did more than 20 brachytherapy applications per month, representing the high burden of disease. 37% (29/79) admitted patients for both intracavitary and interstitial brachytherapy, 22% (17/79) admitted only for interstitial brachytherapy while 27% (21/79) did not admit patients for brachytherapy procedures.

In our data, the respondents who were doing more than 20 applications per month were most commonly using spinal anaesthesia 38%(17/45) in intracavitary brachytherapy.

Of the participants 56% (44/79) also did hybrid brachytherapy application. Most of them used vendor made applicators 43% (40/93) while 9.8% (9/93) did free hand brachytherapy applications.

42% (33/79) of the respondents used transabdominal ultrasound guidance >80% of times and 14% (11/79) used both transabdominal and transrectal USG guidance during application. 6.3% (5/79) used CT guidance for application while 2.5% (2/79) also used MR guidance for application. The majority (91% - 71/79) of the institutions used CT imaging for confirmation of applicator placement while 19% (15/79) used MRI for confirmation of placement 10-30% of times.

Most of the respondents (53%- 53/79) used GEC ESTRO based HRCTV prescription and 44% (44/79) used ICRU point based prescription.

### Analgesia and anesthesia practices

The most common anaesthesia used in intracavitary brachytherapy was spinal anaesthesia alone 21% (21/79) and 39% (45/114) in combination with others as well, followed by General anaesthesia alone 18% (14/79). 4.4% (5/114) were also using oral analgesia for intracavitary application. (Upset plot 1)

In hybrid application also, spinal anaesthesia was most commonly used by 30% (32/106) of participants followed by epidural anesthesia which was used by 20% (21/106) institutes, followed by general anesthesia (13%: 14/106). Remaining responses utilised a combination of 2 or more AS/AG.

In interstitial brachytherapy, 36% (39/109) used spinal anesthesia followed by epidural and general anesthesia. The majority of the participants did not use conscious sedation as it was not preferred by them clinically. 7.7% (7/94) did not do it because of lack of resources and 3.3% (3/94) due to lack of time.

When asked the reason for not using general anesthesia, the majority 25%(27/108) deemed it a clinical preference while 9.3% (10/108) lacked the resources to use GA. During waiting periods i.e. contouring and planning, majority of the participants used oral analgesia (19%) followed by spinal anesthesia, epidural anesthesia and conscious sedation (Upset plot 2) During treatment also oral analgesia was most commonly used (20%: 20/99). At the time of just before the removal, again oral analgesia was most commonly used 32% (27/92) followed by conscious sedation 19% (16/92) and epidural anesthesia 13% (11/92). (Upset plot 3)

On analyzing the description of bivariate distributions, most of the respondents who used epidural anesthesia were from the south while spinal anaesthesia and conscious sedation were most commonly used in the north Indian settings. Of the respondents using oral anaesthesia, most were from the south. All of the centres did more than 20 brachytherapy in a month. Most of the respondents who believed that the intracavitary procedures cause distressing symptoms, utilised epidural anaesthesia followed by spinal and local nerve block. All of the respondents who were utilising local nerve block and conscious sedation during interstitial or hybrid brachytherapy believed that patients had distressing symptoms after the procedure. All these times, the patient was awake during the procedure.

**Upset plot 1.**
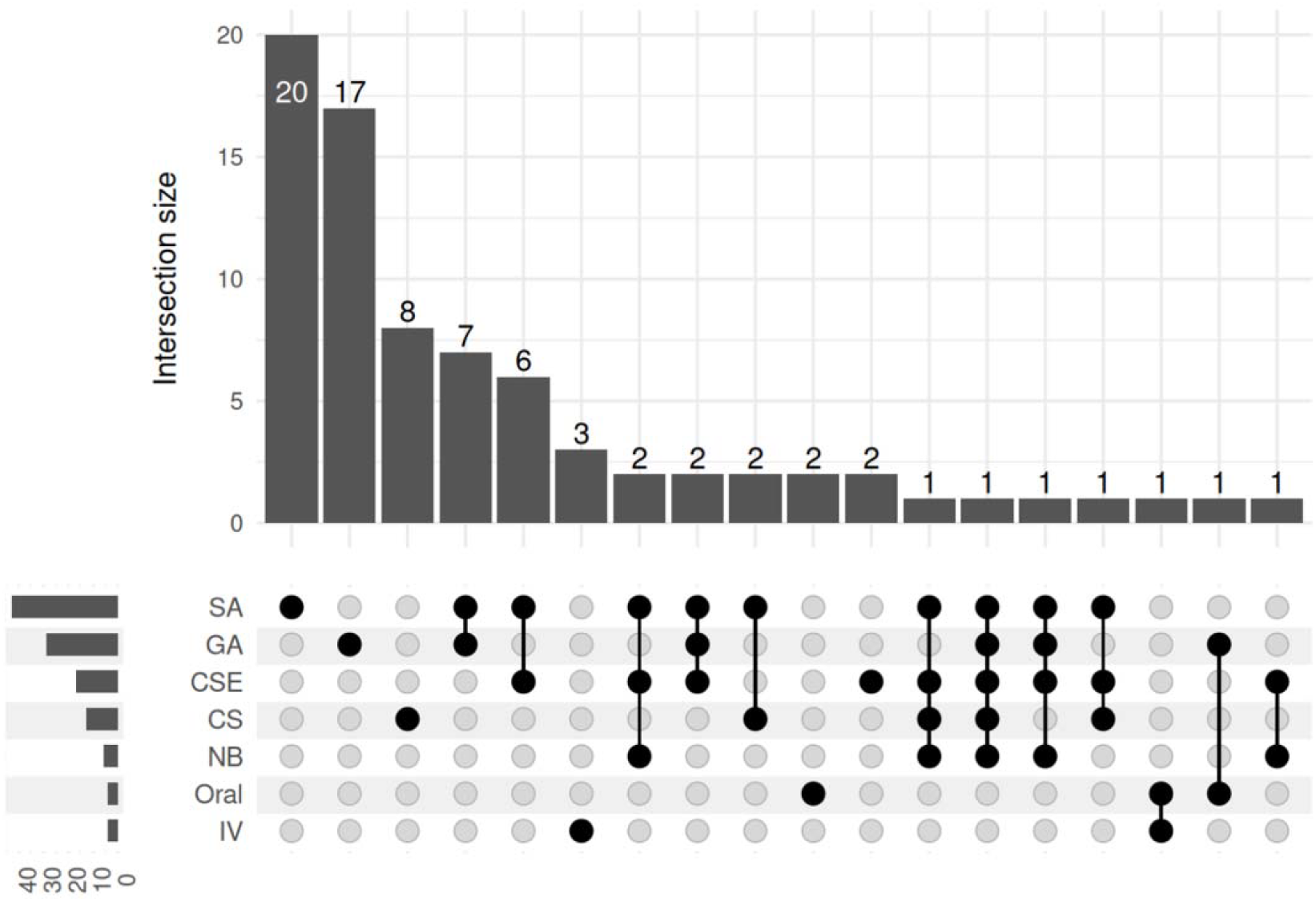
Showing types of AG/AS used during intracavitary brachytherapy application. (Abbreviations: SA: Spinal Anaesthesia, GA: General Anaesthesia, CSE: Combined Spinal and Epidural anaesthesia, CS: Conscious sedation, ND: Nerve block, IV: Intravenous anaesthesia/analgesia.)

**Upset plot 2.**
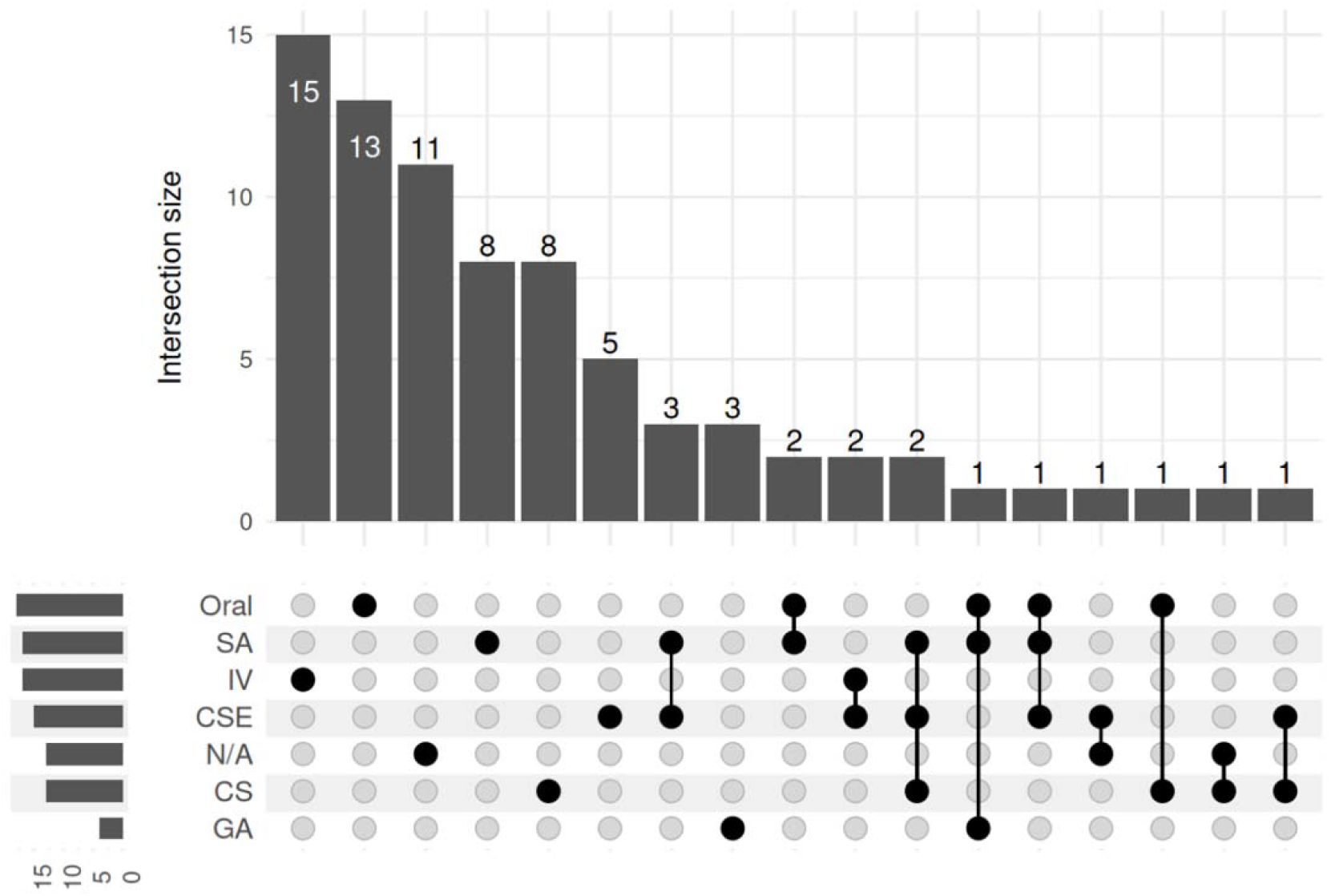
showing types of AG/AS used during intracavitary brachytherapy waiting periods. (Abbreviations: SA: Spinal Anaesthesia, GA: General Anaesthesia, CSE: Combined Spinal and Epidural anaesthesia, CS: Conscious sedation, ND: Nerve block, IV: Intravenous anaesthesia/analgesia, N/A)

**Upset plot 3.**
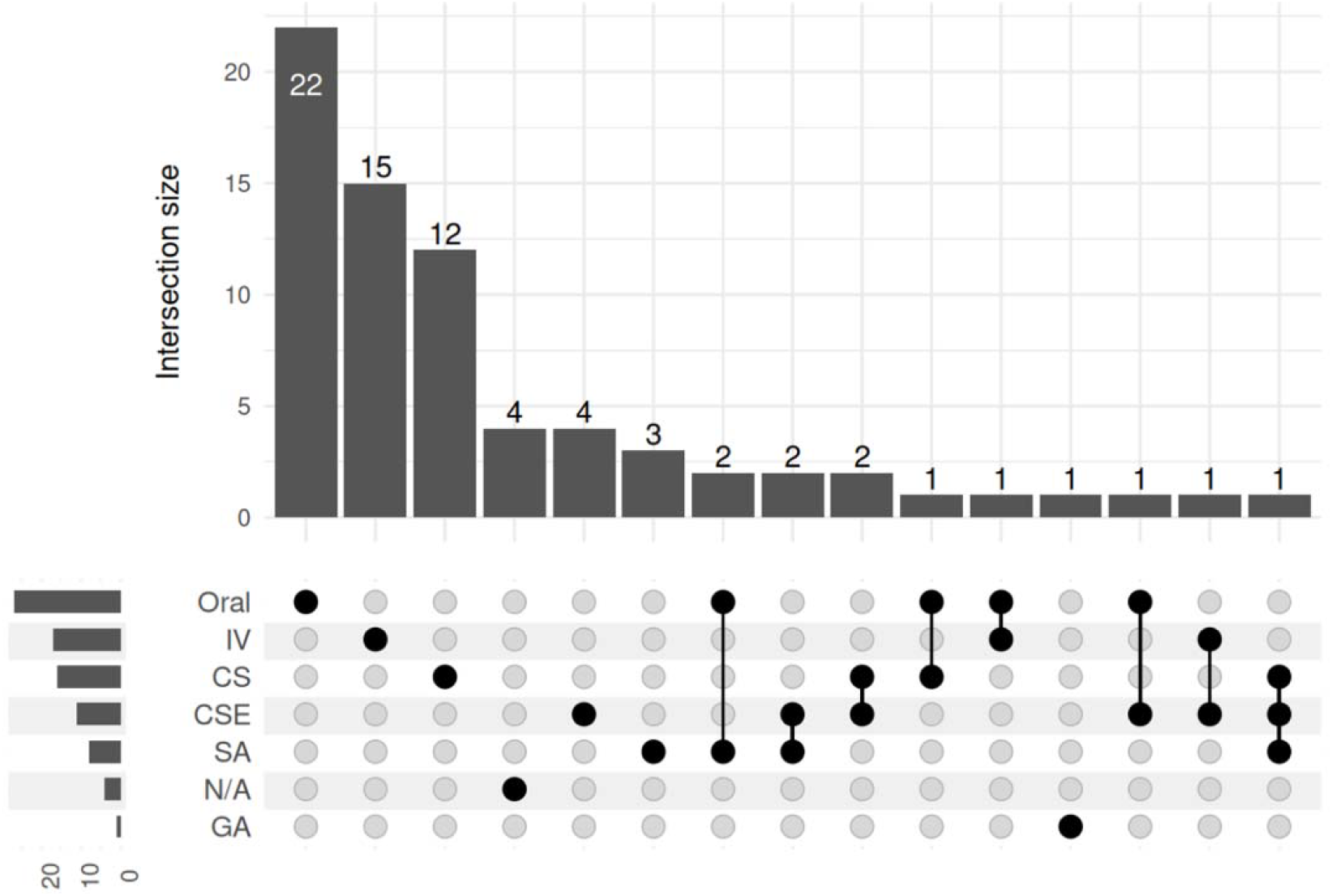
showing types of AG/AS used during intracavitary brachytherapy applicator removal(Abbreviations: SA: Spinal Anaesthesia, GA: General Anaesthesia, CSE: Combined Spinal and Epidural anaesthesia, CS: Conscious sedation, ND: Nerve block, IV: Intravenous anaesthesia/analgesia.)

## Discussion

Brachytherapy in cervical cancer is an inevitable part of treatment, all aspects of which needs to be explored and effort should be made to improve the practice and reduce discomfort to the patient. The first step towards which is knowing how the procedure is being done in highly populated and low middle income country like India. The survey was one of its kind in the country to report the practices of AS/AG in gynaecological brachytherapy. A similar survey wa conducted in the USA^7^ which was modified as per our general practices but also keeping it comparable with the practices in the USA. The sample size which was calculated as per the review of literature was coming out to be 24. Considering the large number of centers in our country, the responses were not restricted. Of the responses we got, 49 institutes were registered with NCG of the total 93 institutes. In contrast to the USA survey, the questionnaire was circulated only with physicians (radiation oncologists) and did not include medical physicists as respondents. The survey in the USA was sent to 90 radiation oncology academic programs out of which they got 41 responses, making the response rate of 46%. As this was the only similar survey, so a response rate of 40% was aimed for with the absolute margin of error being 20%. With this the sample size that was calculated was 24. However for a vast country like India, this would have been a very small sample size. Hence the sample size was not limited. Moreover, in India there is no centralized list of such academic centers. Hence we opted for a convenience sampling strategy. The response rate in our survey if NCG centers alone are considered is 23.8%

The major differences that were noticed when we compared responses with the USA were that the India has a considerably higher case-load of brachytherapy with 38% of respondents doing the procedure for more than 20 patients per month while in the US it is only 7.3% while 61% respondents there did the procedure for up to 5 patients a month. Another stark difference was noted with the type of hybrid applicator used, while it is mostly vendor-made in the USA (80%) in India, it is only 43% highlighting the limited resources in the country. However, even with constrained resources available, Indian physicians are well utilising AS/AG during brachytherapy procedures. Majority 39% (45/114) of the respondents use spinal anaesthesia in application of ICBT. While in the USA the most common AG is GA followed by Conscious sedation (CS). CS has been underutilised in our setting, however it gives sufficient pain control and also opens the window to add analgesia as per individual requirements with patient feedback.^8^ However, it involves administration of sedatives and anxiolytics to provide amnesia and analgesia while maintaining the airway and breathing. This requires certain drugs and monitors and availability of backup in case of respiratory compromise. A lot of these things are not available in all centers in India performing brachytherapy. And thus the underutilisation of CS in our setting.

## Comparison of Brachytherapy Practices: India vs USA (Rivera et al)

## INTRACAVITARY BRACHYTHERAPY

| Parameter | India | USA (Rivera et al) |
| --- | --- | --- |
| No. of Brachytherapy performed in 1 month (>15) | 49.3% | 12.2% |
| Admission for ICA/ISBT | 73.4% (58/79) | 27.5% (11/40) |
| Most common AS/AG | 34% (14/41): General Anesthesia Alone | OA Alone (10% : 4/41) |

## HYBRID BRACHYTHERAPY

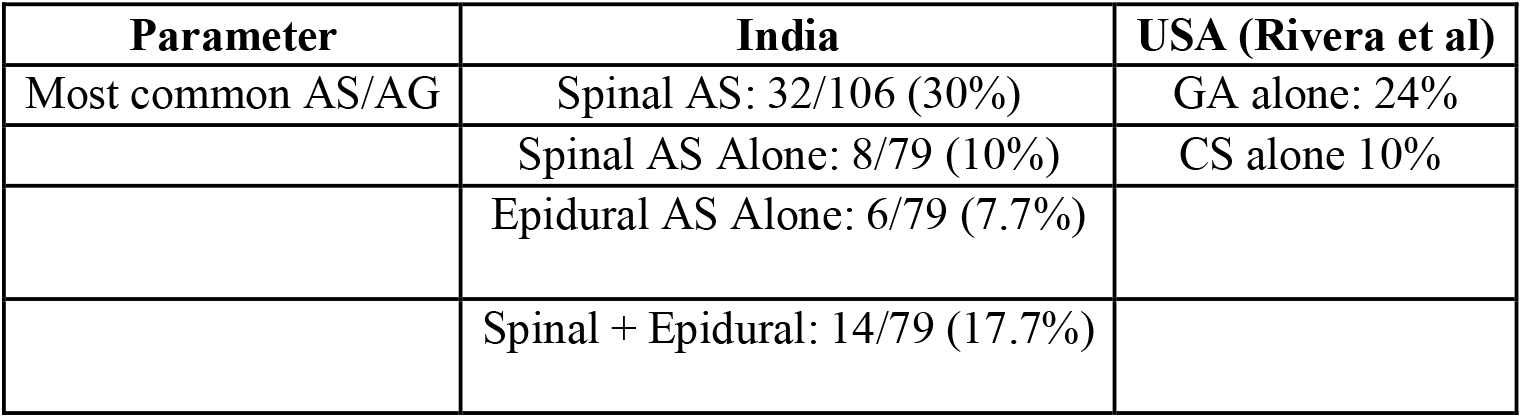

| Parameter | India | USA (Rivera et al) |
| --- | --- | --- |
| Most common AS/AG | Spinal AS: 32/106 (30%) | GA alone: 24% |
|  | Spinal AS Alone: 8/79 (10%) | CS alone 10% |
|  | Epidural AS Alone: 6/79 (7.7%) |  |
|  | Spinal + Epidural: 14/79 (17.7%) |  |

## EMOTIONAL DISTRESS

| Parameter | India | USA (Rivera et al) |
| --- | --- | --- |
| Emotional Distress - Yes | 39/79 (49%) | 30/41 (73%) |
| Measures to alleviate emotional distress - Yes | 35 (44%) | 28/41 (68%) |

From the data available from the west, it has been reported that there is severe pain associated with intracavitary applicator insertion, with variability in analgesia and anesthesia practices across institutions.^4-7^ Even with intravenous patient controlled regimens, 46% women reports unacceptable pain scores. Better pain control was reported with intermittent epidural bolus with patient controlled epidural analgesia.^19^ A meta-analysis by Petit et al reported reduced frequency of administration of rescue analgesics with neuraxial anaesthesia as compared to general anaesthesia.^2^ However multimodal approach to pain control might be required for better comfort of the patient.^9^

An observational study done in Japan including 100 brachytherapy sessions of 27 patients who were given NSAID suppositories after assessing pain scores, 29.6% required anesthesia over suppositories. The benefit is that if tolerated well, does not always require involvement of an anaesthesiologist and airway management.^10^ A study done in Ottawa, Canada using conscious sedation as a primary anesthesia, observed that most patients were able to tolerate the whole procedure with only some incidental moderate to severe pain when the applicator was manipulated. It was also noted that bolus before critical parts of procedure like application and removal improved pain tolerance during procedure.^8^ The benefits with conscious sedation is that it reduced both induction and recovery time and also does not require admission allowing the procedure done on OPD basis. However, the use of conscious sedation is rare in Indian settings, mainly due to physician preference and possibly due to lesser experience with the same.

Recent advances include ultrasound-guided paracervical and sacral nerve blocks, which significantly reduce procedural pain scores while maintaining hemodynamic stability.^11 7^ Comparative studies further suggest that spinal anesthesia may offer superior pain control over general anesthesia or sedation during MRI-based insertions, enhancing both patient comfort and procedural reproducibility.^5^ Nevertheless, anesthesia choices often depend on institutional logistics, operator preferences, and patient comorbidities rather than formal protocols.^7 18^

49% of the respondents also believed that the whole procedure causes distress to the patient. While 44% also incorporate measures to deal with it. The details of the practices to reduce stress of the patient were not covered in the survey.

There is evidence highlighting the pain and distress a patient goes through during this treatment. No data however emphasises on the perception of the physicians regarding the same and if they incorporate any strategy or special measures to reduce the same. When we tried to enquire about the same we realised that less than half of the responders believe that the patient goes through emotional distress and even fewer (44%) integrate any steps to reduce that. This area is unsought and usually goes unnoticed, but requires more attention along with efforts to reduce pain caused during procedures.

Pain and symptom monitoring remain underutilized. We need to serially assess patients across multiple fractions to guide individualized analgesia adjustments and identify patients needing psychological support.^19 20^ The role of integrative interventions, such as physiotherapy, yoga, and educational programs, has shown promise in reducing perceived stress and improving overall experience.^16^

An often-overlooked issue is the influence of anesthesia modality on treatment quality and dosimetry. Studies have shown that inadequate anesthesia may compromise applicator placement, reproducibility, and dwell time optimization, ultimately affecting dose distribution and treatment efficacy.^14 18^ This underscores the need for standardized anesthesia protocols that are not only safe but also optimize brachytherapy delivery.

In conclusion, as brachytherapy techniques continue to evolve, there is a need to combine clinical precision with patient-centered care. Pain control and psychological well-being should be integral components of brachytherapy protocols. Future efforts must focus on standardizing analgesia practices, enhancing interdisciplinary collaboration and incorporating routine patient feedback to ensure holistic, high-quality care for women with cervical cancer.

## Data Availability

All data produced in the present study are available upon reasonable request to the authors

## Limitations

The survey had relatively lesser responses than expected. The reasons include unavailability of institutional programmes that could be contacted through institutional channels leading to lesser responses. The National Cancer Grid also does not provide online information for contacting the institutes registered with it. There still are multiple institutes that are doing oncological practices but are not registered with the NCG.

## Declarations

No conflict of interests.

